# Regional distribution of anoxic brain injury after cardiac arrest: clinical and electrographic correlates

**DOI:** 10.1101/2021.05.14.21257192

**Authors:** Samuel B. Snider, David Fischer, Morgan E. McKeown, Alexander L. Cohen, Frederic L.W.V.J. Schaper, Edilberto Amorim, Michael D. Fox, Benjamin Scirica, Matthew B. Bevers, Jong W. Lee

## Abstract

**Introduction:** Disorders of consciousness, EEG background suppression and epileptic seizures are associated with poor outcome after cardiac arrest. The underlying patterns of anoxic brain injury associated with each remain unknown. Our objective was to identify the distribution of anoxic brain injury after cardiac arrest, as measured with diffusion MRI, and to define the regional correlates of disorders of consciousness, EEG background suppression, and seizures.

**Methods:** We analyzed patients from a prospectively-maintained, single-center database of unresponsive patients who underwent diffusion-weighted MRI following cardiac arrest (*n* = 204). We classified each patient based on recovery of consciousness (command-following) before discharge, the most continuous EEG background (burst suppression versus continuous), and the presence or absence of seizures. Anoxic brain injury was measured using the apparent diffusion coefficient (ADC) signal. We identified abnormalities in ADC relative to control subjects without cardiac arrest (*n* = 48) and used voxel lesion symptom mapping to identify regional associations with disorders of consciousness, EEG background suppression, and seizures. We then used a bootstrapped lasso regression procedure to identify robust, multivariate regional associations with each clinical and EEG variable. Finally, using area under receiver operating characteristic curves, we then compared the classification ability of the strongest regional associations to that of brain-wide summary measures.

**Results:** Compared to control subjects, cardiac arrest patients demonstrated a reduction in the ADC signal that was most significant in the occipital lobes. Disorders of consciousness were associated with reduced ADC most prominently in the occipital lobes, but also in the basal ganglia, medial thalamus and cerebellar nuclei. Regional injury more accurately classified patients with disorders of consciousness than whole-brain injury. Background suppression mapped to a similar set of brain regions, but regional injury could no better classify patients than whole-brain measures. Seizures were less common in patients with more severe anoxic injury, particularly in those with injury to the lateral temporal white matter.

**Discussion:** Anoxic brain injury was most prevalent in posterior cerebral regions, and this regional pattern of injury was a better predictor of disorders of consciousness than whole-brain injury measures. EEG background suppression lacked a specific regional association, but patients with injury to the temporal lobe were less likely to have seizures. Collectively, our results suggest that the regional pattern of anoxic brain injury is relevant to the clinical and electrographic sequelae of cardiac arrest and may hold importance for prognosis.

## Introduction

Neuropathologic studies in animals^1^ and humans^2, 3^ have identified numerous brain regions susceptible to anoxic injury, but a map of brain regions affected by cardiac arrest does not yet exist. Furthermore, although disorders of consciousness (DoC)^4, 5^, EEG background suppression^6, 7^ and seizures^7-10^ predict poor neurological outcomes after cardiac arrest ^3, 11, 12^, the regional patterns of anoxic brain injury (ABI) associated with each are unknown^3, 13^.

Diffusion MRI is the most sensitive imaging modality for the clinical characterization of anoxic brain injury (ABI)^14-19^. In animal models of anoxic brain injury, reductions in the apparent diffusion coefficient (ADC) are associated with the histopathological volume of infarcted brain tissue^20^. Previous work has demonstrated that greater ADC reductions across the whole brain or cortex are associated with poorer outcomes^21^.

Furthermore, it remains unclear whether common clinical and electrographic abnormalities following cardiac arrest – such as DoC, a suppressed EEG background, and post-anoxic seizures – reflect the overall severity of ABI, as measured with whole-brain or whole-cortex ADC values, or injury to specific structures^13^. Investigating the associations between regional anoxic injury and the clinical and electrographic sequalae of cardiac arrest may help identify specific brain circuits responsible for their production. Furthermore, given an increasingly multi-modal approach to prognostication after cardiac arrest^22-24^, an improved understanding of the associations between modalities will facilitate interpretation of discrepant results.

Here, we use diffusion MRI from a large clinical cohort to define the anatomic distribution of ABI after cardiac arrest and to identify associations between patterns of injury and common clinical and electrographic abnormalities.

## Materials and methods

### Cardiac arrest cohort

This study was approved by the Mass General Brigham institutional review board (IRB). We identified patients from a prospectively-maintained registry of 573 patients with in- or out-of-hospital cardiac arrest at Brigham and Women’s Hospital between 2009 and 2020. Criteria for inclusion in the registry included age > 18 years old and the absence of command-following on the first assessment after return of spontaneous circulation. 206 of these patients had a diffusion-weighted MRI within 14 days of cardiac arrest, with 204 completing the processing pipeline outlined below.

### Clinical covariates

Basic demographic information, cardiac rhythm, the use of targeted temperature management (TTM), and time between cardiac arrest and MRI acquisition were abstracted from the electronic medical record (Table 1). DoC was defined as the absence of command-following documented at any time after cardiac arrest in the medical record. Patients with missing data were excluded from the relevant analysis.

**Table 1.** cohort characteristics.

|  | **Cardiac Arrest**<br>**(n = 204)** | **Controls**<br>**(n = 48)** | **P value** |
| --- | --- | --- | --- |
| Mean Age (95% CI) | **55 (53, 58)** | **51 (47, 55)** | **0.04** |
| **Male Sex (%)** | **131 (64%)** | **13 (27%)** | **< 0.001** |
| **White Ethnicity (%)** | **130 (64%)** | **36 (82%)** | **0.04** |
| MRI Field Strength (% 1.5T) | 145 (71%) | 27 (59%) | 0.2 |
| MRI manufacturer (% Siemens) | 177 (87%) | 34 (77%) | 0.2 |
| Shockable Cardiac Rhythm (%) | 84 (41%) |  |  |
| Received TTM (%) | 176 (86%) |  |  |
| Followed Commands Prior to Discharge (%) | 80 (39%) |  |  |
| Died Prior to Discharge (%) | 116 (58%) |  |  |
| EEG (%) | 193 (95%) |  |  |
| Mean Days Between Arrest and MRI (95% CI) | 5.0 (4.6, 5.3) |  |  |

### EEG background continuity

193 patients also had continuous (*n* = 186) or routine (*n* = 7) EEG data. EEG data were recorded using Natus XLTEK system (Pleasanton, CA) according to the international 10-20 system and interpreted using the American Clinical Neurophysiology Society (ACNS) critical care EEG terminology^25^. Beginning in February 2013, background continuity of each epoch was prospectively categorized^26^ by a board-certified epileptologist as continuous, nearly-continuous (suppression [< 10 uV] < 10% of record), discontinuous (suppression 10-49% of record), burst-suppression (suppression 50-90% of record) and overall suppression (suppression > 90% of record). The presence or absence of intravenous sedating medications during each epoch was also recorded. For records predating 2013, background continuity and sedating medications were manually abstracted from EEG reports. Records with ambiguous documentation were excluded. A single continuity value was assigned to each 24-hour EEG epoch, reflecting the highest continuity level within that epoch. Each patient was classified by the most continuous EEG background observed after cardiac arrest. For further analyses, the best-achieved continuity was then binarized on the absence or presence of EEG background suppression (a category including burst suppression and overall suppression).

### Seizures

The presence of seizures was determined per EEG epoch as per the modified Young criteria for non-convulsive electrographic seizures^27^ or Salzburg criteria for status epilepticus^28^. Patients were dichotomized as either having or not having seizures after cardiac arrest.

### Image processing and quality control

When multiple MRIs were performed, only the first MRI was included in the analysis. Patients’ clinically-acquired T1 and diffusion sequences were used. Scans were acquired on several clinical magnets over the course of the study, including Siemen’s Verio (3T), TrioTrim (3T), and Aera (1.5T), and GE Signa (1.5T) and Discovery (3T). Parameters of the diffusion acquisition varied but included b=0, 500 and 1000 seconds /mm^2^, 1.5 × 1.5 × 6mm voxel size, and diffusion encoding along the principal axes. T1 weighted acquisitions included either sagittal and axial pre-contrast volumes (voxel size ∼ .5 × .5 × 5mm) or an MPRAGE volume (voxel size 1 × 1 × 1 mm).

To register the ADC maps into a standardized space, we performed a brain extraction (FSL, bet2) and concatenated linear, non-linear, and diffeomorphic registrations (ANTs: antsRegistrationSyNQuick) between the diffusion b0 volume, the T1 weighted MRI, and MNI152 T1 space. This was accomplished using code from https://github.com/bchcohenlab/bids_lesion_code. When a b0 volume was unavailable, the ADC map was used for cross-modality registration. If a T1 weighted isotropic MPRAGE sequence was not available, clinical axial and sagittal T1 volumes were combined using NiftyMIC (https://github.com/gift-surg/NiftyMIC) and resliced into an isotropic volume for registration. All registrations were then manually inspected using slicesdir (FSL 6.0.1). Two patients’ MRIs were excluded due to grossly inaccurate registrations by visual inspection.

To minimize signal contribution from artifact and cerebrospinal fluid, we thresholded the ADC maps between 200 and 1200 × 10^−6^ mm^2^/s, as done previously^15-17^. The above transformations were then applied to the thresholded ADC maps to bring them into MNI space for group-level comparisons.

### Non cardiac-arrest controls

To identify a group of patients unlikely to have significant brain pathology, we screened our hospital system electronic medical record for MRI reports containing the text string ‘migraine headaches’ between 2000 and 2020. From this list of over 6000 patients, we randomly selected 150. Of those, 57 were from our specific institution and therefore comparable to the cohort of patients with cardiac arrest; these were processed in the same manner as above. After exclusion of 9 patients missing diffusion or T1 sequences, or with grossly inaccurate registrations, 48 were included for further analysis (Table 1, Controls).

### Between-group differences in baseline characteristics

Differences in demographic, clinical, and radiologic characteristics between subjects with or without cardiac arrest, with or without DoC, with varying EEG background continuity, and with or without seizures were calculated using one-way ANOVA (continuous variable, 3 groups), 2 sample t test (continuous variable, 2 groups) or Chi-Squared tests (binary variable).

### Voxel lesion symptom mapping (VLSM)

VLSM was conducted using randomise (FSL 6.0.1). Given that there is no agreed upon ADC threshold to predict tissue infarction after cardiac arrest^14-16^ or ischemic stroke^29, 30^, we treated the ADC signal as a continuous variable. To mitigate any possible effects of registration variability and sulcal volume differences between cardiac arrests and controls, we excluded, from all subjects, voxels with ADC values of 0 (indicating a pre-threshold ADC value of < 200 or > 1200) in >20% of subjects (Supplementary Figure 1A). Contrasts were generated using 2-sample T tests with or without covariates (FSL, randomise^31^), with 2000 permutations and threshold free cluster enhancement (TFCE)^32^. Because quantitative diffusion metrics, like ADC, are known to vary by age^33^, time from injury^34^, MRI manufacturer^35^ and field strength^36^, we conducted additional analyses controlling for these values as nuisance covariates, mean-centering the continuous variables. To ensure the results were not dependent on the processing strategy, we replicated the primary analyses after binarizing each subject’s ADC map at previously-used^15, 16^ ADC thresholds of 550 or 650 × 10^−6^ mm^2^/s and after normalizing each subject’s ADC map to the mean value within the brainstem, a region rarely affected by ABI^19^.

### Region of interest ADC measurement

Using the previously described transformations, we transformed binary, MNI-space *a priori* ROIs onto each subject’s thresholded ADC map and computed the robust (non-zero) mean ADC value within each ROI. Grey-matter ROIs included bilateral frontal, temporal, insular, parietal and occipital lobes and cerebellum (MNI Structural Atlas^37^), bilateral basal ganglia (combined caudate, putamen, and pallidum), bilateral thalami and brainstem (Harvard-Oxford Subcortical Atlas^38^). White matter ROIs included all bilateral and binarized tracts in the Johns Hopkins University White Matter Atlas^39^ as well as the corpus callosum (Julich Atlas^40^).

### Multivariate regional associations with outcome

We observed substantial collinearity between mean ADC measurements of different brain regions (Pearson R: 0.30 to 0.99, Supplementary Figure 2). Combining ADC measurements from multiple ROIs into a single regression model yielded unstable effect estimates, precluding the identification of independent regional predictors. We therefore employed lasso regression, a technique that penalizes models with larger numbers of coefficients and is well-suited to winnowing multiple colinear predictor variables (R package: ‘glmnet’^41^). Modeling binary outcomes, (DoC, background suppression, or seizures), we included each anatomical ROI as a candidate predictor variable. All variables were mean-centered and scaled to unit variance. We computed the receiver operating characteristic (ROC) curve (R packages: ‘pROC’^42^) for the model, as well as for the whole-brain and whole-cortex mean ADC values. We then compared the area under the receiver operating characteristic curve (AUROC) of the lasso regression model to that of whole-brain and whole-cortex ADC using Delong’s test^43^.

To ensure these results were not driven by outliers, we performed a repeated, stratified cross-validation procedure, iteratively training a lasso regression model with anatomical ROIs in 50% of the data and testing in the remaining 50%, maintaining the ratio of outcomes between samples (R package: ‘caret’). For each of 1000 iterations, we computed the AUROCs for the lasso regression, mean whole-brain ADC and mean whole-cortex ADC in the test sample. We then reported the percentage of times that the model AUROC was larger than the whole-brain and whole-cortex AUROC.

### Data availability

Anonymized clinical and MRI data will be made available to investigators upon reasonable request. We plan to upload the full set of anonymized MRI volumes to openneuro.org pending IRB approval.

## Results

### Distribution of anoxic brain injury after cardiac arrest

Characteristics of the 204 patients who were unresponsive after cardiac arrest and underwent diffusion MRI are provided in Table 1. Control subjects were slightly younger, had a higher proportion of women, and a higher proportion of subjects identifying as white (Table 1, *P* < 0.05). To determine the brain regions most affected by anoxia, we compared the voxel-wise distributions of ADC values between patients who did (*n* = 204 Figure 1A) and did not (*n* = 48, Figure 1B) sustain a cardiac arrest. Compared to control subjects, cardiac arrest patients had lower ADC values across multiple cortical regions, most prominently in the occipital lobes (*P*_TFCE_ < 0.05, Figure 1C). An ROI-based analysis confirmed a significant region by group interaction (*F*_21,252_ = 2.0, *P* = 0.005), with the occipital lobes showing largest ADC difference between groups (*T* = 4.6). The set of identified regions remained consistent after controlling for possible sources of confounding including demographics (Supplementary Figure 3A), MRI field strength (1.5 vs 3T, Supplementary Figure 3B), and MRI manufacturer (Siemens vs GE, Supplementary Figure 3C). The areas of injury identified after binarizing the ADC maps with previous thresholds^15^ (550, 650 × 10^−6^ Supplementary Figure 3D-E) were broader but located in similar regions.

**Figure 1.**
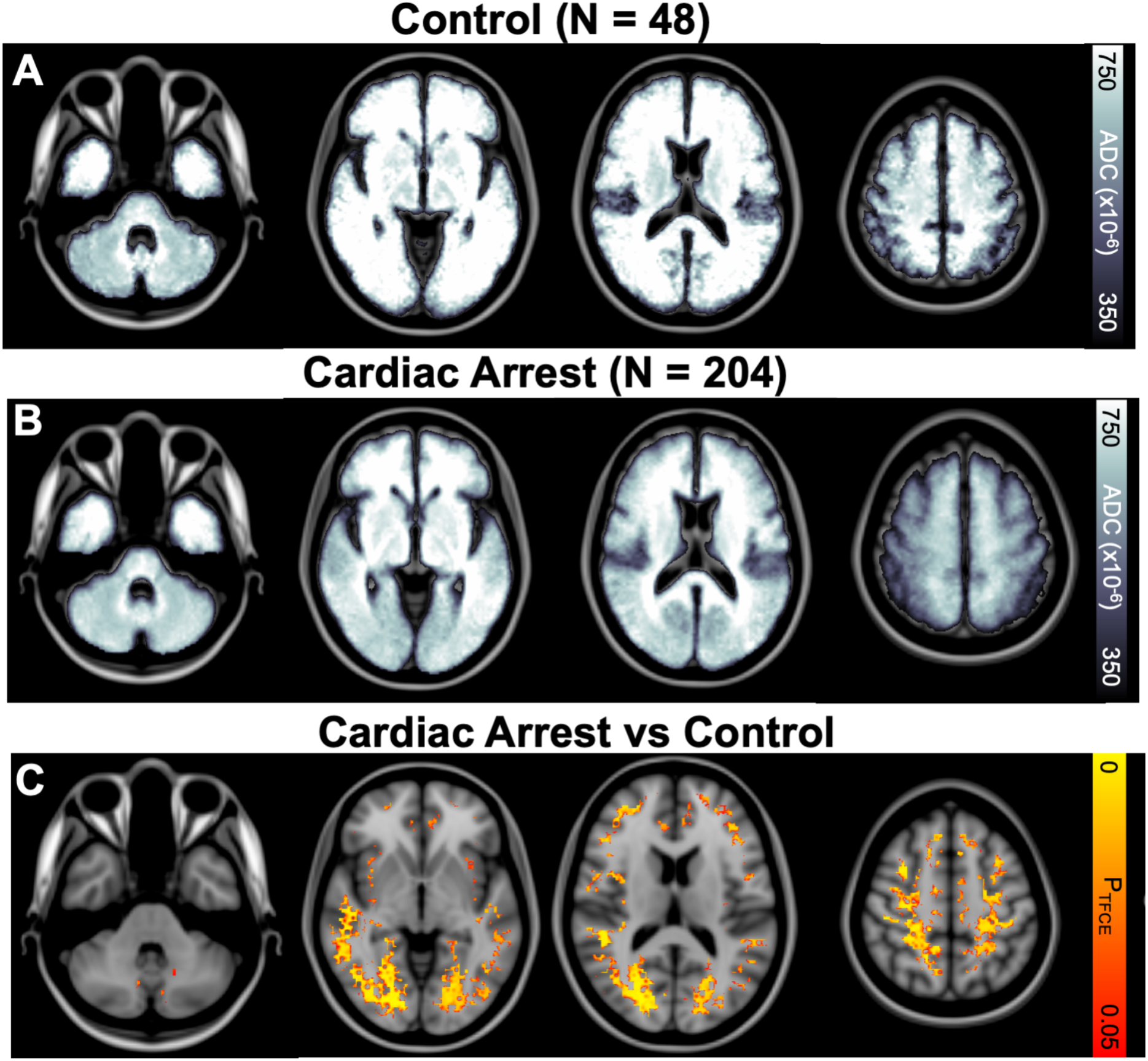
Distribution of anoxic brain injury after cardiac arrest. **(A)** Mean ADC map for control subjects. **(B)** Mean ADC map for all patients with cardiac arrest. **(C)** Voxels with significantly lower ADC in cardiac arrest patients as compared to controls are localized at the grey-white junction, most prominent occipitally. Map corrected for multiple comparisons using threshold free cluster enhancement (TFCE).

### DoC: distribution of injury

The cardiac arrest cohort contained a mix of patients with (CARecovered, 39%) and without (CA_DoC_, 61%) recovery of consciousness, as measured by command-following (Table 2). We sought to determine which brain regions were specifically lesioned in CA_DoC_ compared with CA_Recovered_ patients. Compared to CA_Recovered_ patients (Figure 2A), CA_DoC_ patients (Figure 2B) had lower ADC values across a range of subcortical and cortical regions, most prominently in the occipital lobes (*P*_TFCE_ < 0.05, Figure 2C). An ROI-based analysis again confirmed this interpretation, with a region by group interaction (*F*_21,203_ = 4.7, *P* < 0.001) and the largest between-group ADC difference in the occipital lobes (*T* = 9.4). Though CA_DoC_ patients were older than the CA_Recovered_ patients (Table 2, *P* < 0.001), controlling for age did not alter the resulting maps (Supplementary Figure 4). Whole-brain ADC differed between CA_DoC_, CA_Recovered_, and controls (*F*_2,250_ = 20, *P* < 0.001, Figure 2D), with ADC higher in CA_Recovered_ compared to controls (*T* = 3.7, *P* < 0.001, Figure 2D and Supplementary Figure 5).

**Table 2.** Baseline characteristics in cardiac arrest patients with or without DoC.

|  | **CA<sub>DoC</sub>**<br>**(n = 123)** | **CA<sub>Recovered</sub>**<br>**(n = 80)** | **P value** |
| --- | --- | --- | --- |
| **Mean Age (95% CI)** | **58 (55, 61)** | **52 (48, 55)** | **< 0.0001** |
| Male Sex (%) | 127 (63%) | 57 (71%) | 0.1 |
| White Ethnicity (%) | 79 (64%) | 50 (63%) | 0.9 |
| MRI Field Strength (% 1.5T) | 91 (74%) | 53 (66%) | 0.3 |
| MRI manufacturer (% Siemens) | 108 (88%) | 68 (85%) | 0.7 |
| **Shockable Cardiac Rhythm (%)** | **31 (25%)** | **52 (65%)** | **< 0.0001** |
| Received TTM (%) | 109 (89%) | 66 (83%) | 0.3 |
| Mean Days Between Arrest and MRI (95% CI) | 4.7 (4.3, 5.1) | 5.3 (4.8, 5.9) | 0.07 |

**Figure 2.**
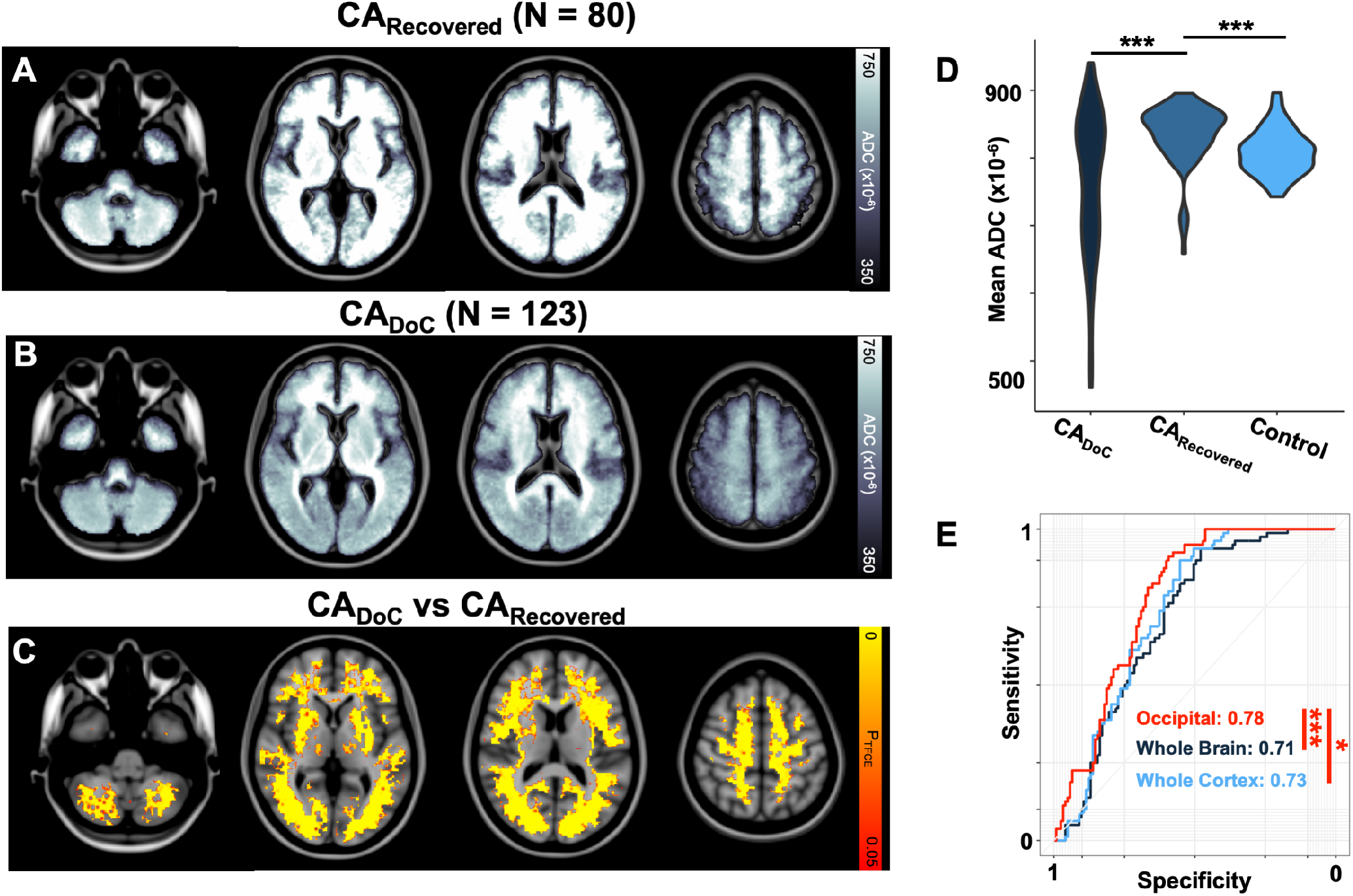
DoC after cardiac arrest is associated with occipitally predominant injury. Group-wide mean ADC maps for subjects with cardiac arrest who did (CARecovered, *n* = 80, **A**) or did not (CA_DoC_, *n* = 123, **B**) recover consciousness after cardiac arrest show areas of different ADC signal between groups. **(C)** Cortical (most prominent occipitally), putamen, globus pallidus, medial thalamus, and cerebellar voxels have significantly lower ADC in CA_DoC_ as compared to CA_Recovered_ *P*_TFCE_ < 0.05. The distribution of whole-brain mean ADC across study groups **(D)**, illustrates the high variance in CA_DoC_ and the elevated mean ADC in CA_Recovered_ relative to controls. **(E)** Area under the receiver operating characteristic curves (AUROC) demonstrate that occipital ADC (red), better classifies subjects with DoC than whole-brain (dark blue), or whole-cortex (light blue) ADC. Area under ROC curves (AUROC) values are listed next to the legend. *** = *P* < 0.0005, ** = *P* < 0.005, * = *P* < 0.05.

### DoC: global versus regional injury

We next sought to determine which set of brain regions were independently associated with recovery of consciousness after cardiac arrest, and whether global or regional injury was a stronger predictor of DoC. To identify the strongest independent regional associations with DoC, we ran a penalized (lasso) regression on the set of all anatomical ROIs, finding that only the occipital ADC had a nonzero coefficient. The AUROC for this occipital-only model (0.78 95% CI [0.71, 0.84]) was significantly greater than that of whole-brain (0.71 [0.64, 0.78], Delong’s Z: 3.6, *P* = 0.0003) or whole-cortex ADC (0.73 [0.67, 0.80], *Z*: 2.7, *P* = 0.008, Figure 2E). Confirming the robustness of these results within this dataset, an ROI-based lasso regression model had a greater AUROC than whole-brain and whole-cortex ADC in 99.5% of 1000 split-half iterations. The occipital ADC was included in 100% of models, and the brainstem and anterior thalamic radiation ADC were included as covariates in 1% and 0.1% of models, respectively.

### EEG background suppression

Characteristics of patients with different EEG background continuities (*n* = 175) are shown in Table 3, and temporal trends in EEG background continuity are displayed in Sup. Figure 6. The prevalence of background suppression decreased by whole-brain ADC tertile (Sup. Figure 7A). Background suppression had a positive predictive value of 65% for overt anoxic brain injury (whole-brain ADC value in the lowest tertile). All the brain regions lesioned in CA_Coma_ patients (Supplementary Figure 1B), were also associated with the presence of background suppression (*P*_TFCE_ < 0.05, Figure 3A), even after adding the elapsed time between arrest and MRI as a nuisance covariate (Supplementary Figure 8A). The AUROC for the lasso regression model (0.81 [0.74, 0.88]), with occipital and cerebellar ADC as its only nonzero coefficients, was no different from that achieved with whole-brain (0.79 [0.71, 0.86] *Z* = 1.7, *P* = 0.1) or whole-cortex ADC (0.80 [0.72, 0.87], *Z* = 1.2, *P* = 0.2, Figure 3B). In a robustness analysis, the lasso regressions included a range of anatomical ROIs (Supplementary Figure 8B) and the model-based AUROC was greater than whole-brain and whole-cortex ADC in only 56% of 1000 split-half iterations.

**Table 3:** Baseline Characteristics across EEG Background Continuities.

|  | **Continuous/Nearly-continuous**<br>**(n = 101)** | **Discontinuous**<br>**(n = 17)** | **Background Suppression**<br>**(n = 57)** | **P value** |
| --- | --- | --- | --- | --- |
| **Mean Age (95% CI)** | **53 (50, 56)** | **55 (45, 65)** | **59 (55, 64)** | **0.03** |
| Male Sex (%) | 63 (62%) | 13 (77%) | 35 (61%) | 0.5 |
| White Ethnicity (%) | 68 (67%) | 8 (47%) | 36 (63%) | 0.4 |
| **Shockable Cardiac Rhythm (%)** | **48 (48%)** | **5 (29%)** | **14 (25%)** | **0.01** |
| Received TTM (%) | 81 (80%) | 15 (88%) | 55 (93%) | 0.09 |
| No sedation (%) | 9 (9%) | 3 (18%) | 8 (14%) | 0.4 |
| **Mean Days Between Arrest and MRI (95% CI)** | **5 (5, 6)** | **4 (4, 5)** | **4 (4, 5)** | **0.02** |
| **Mean Whole-Brain ADC (95% CI)** | **825 (813,836)** | **817 (780,854)** | **719 (689,749)** | **< 0.001** |

**Figure 3.**
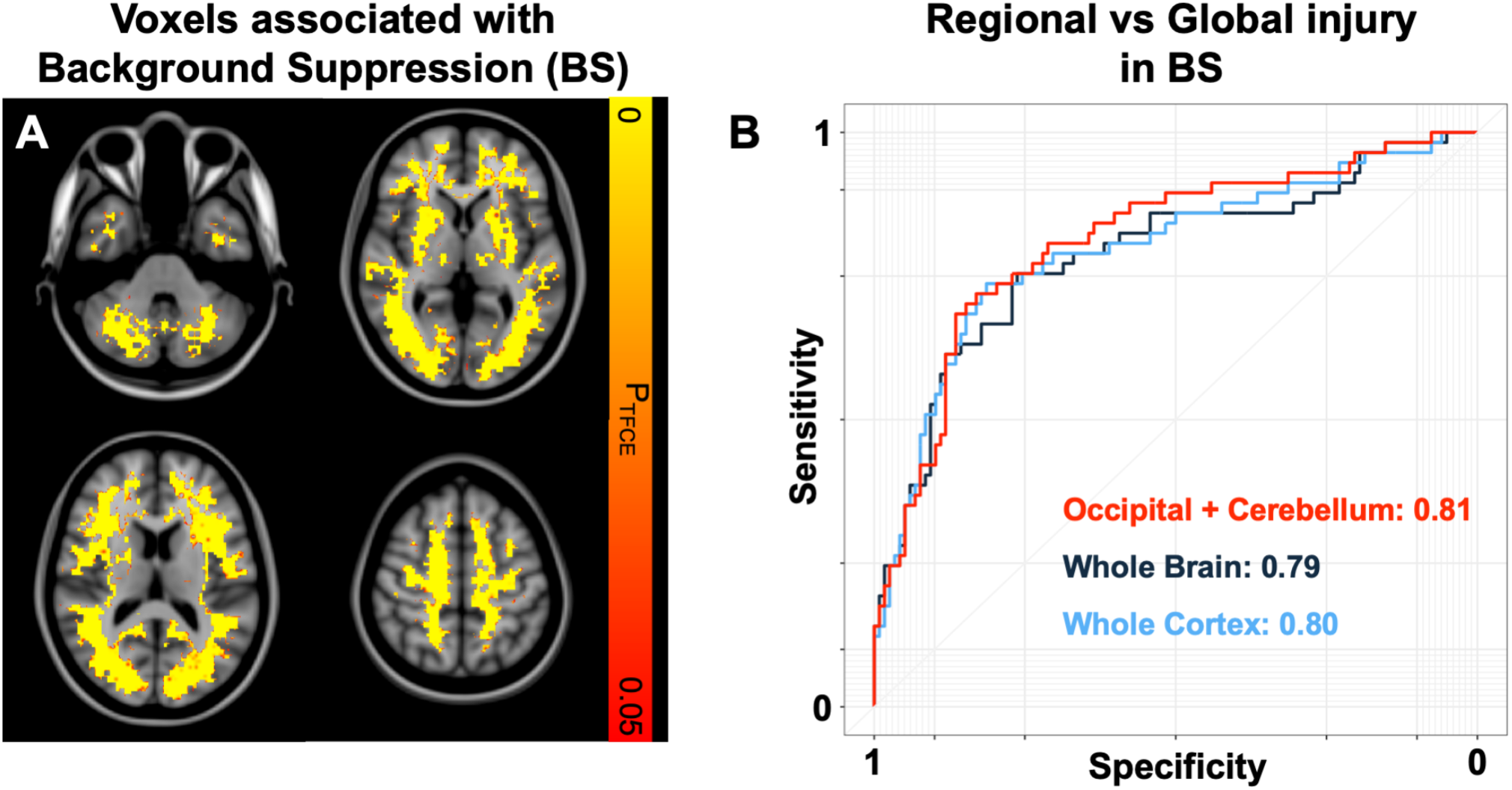
Anoxic brain injury severity is associated with background suppression after cardiac arrest. **(A)** TFCE-corrected map of brain voxels with lower ADC in patients with as compared to without persistent background suppression (BS) appears similar to the map of voxels associated with DoC. **(B)** ROC curves comparing the ability of a lasso regression model including cerebellar and occipital ADC (red) to classify patients with versus without BS to whole-brain (dark blue) and whole-cortex (light blue) ADC. AUROC values are listed next to the legend. The occipital/cerebellar model could no better classify BS patients than whole-brain or whole-cortex ADC.

### Seizures: distribution of injury

Baseline characteristics of patients with (*n* = 35) and without (*n* = 158) seizures are shown in Table 4. Despite no other demographic or clinical differences, whole-brain ADC was lower in patients without (mean 783, 95% CI: [767, 799]) as compared to with (817 [793, 842]) seizures (*P* < 0.05, Table 3, Figure 4A). Seizure incidence decreased non-significantly with each decreasing ADC tertile (*X*^2^_(2,193)_ = 3.6, *P* = 0.2, Supplementary Figure 7B). The ADC signal within lateral temporal white matter showed the strongest (inverse) association with seizures (Figure 4B, *P*_TFCE_ < 0.05).

**Table 4:**
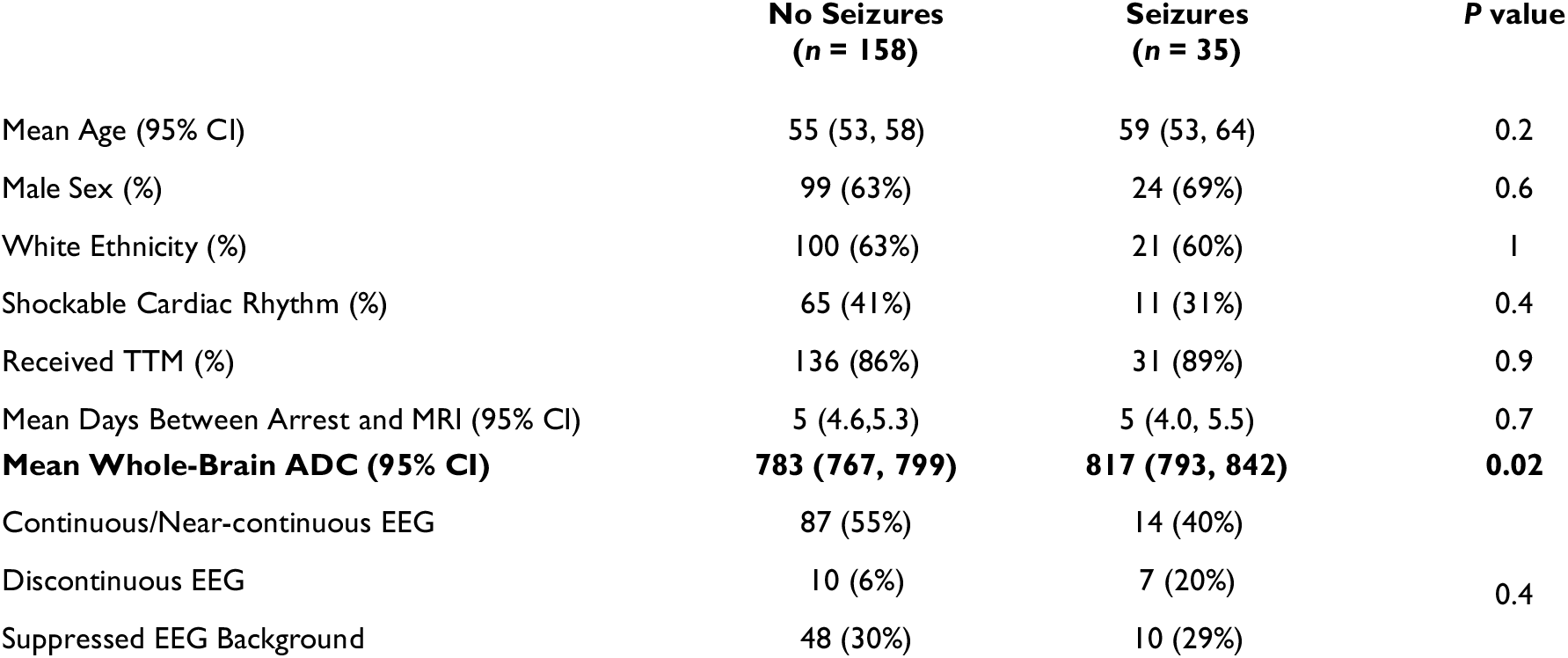
Baseline characteristics in patients with and without seizures.

**Figure 4:**
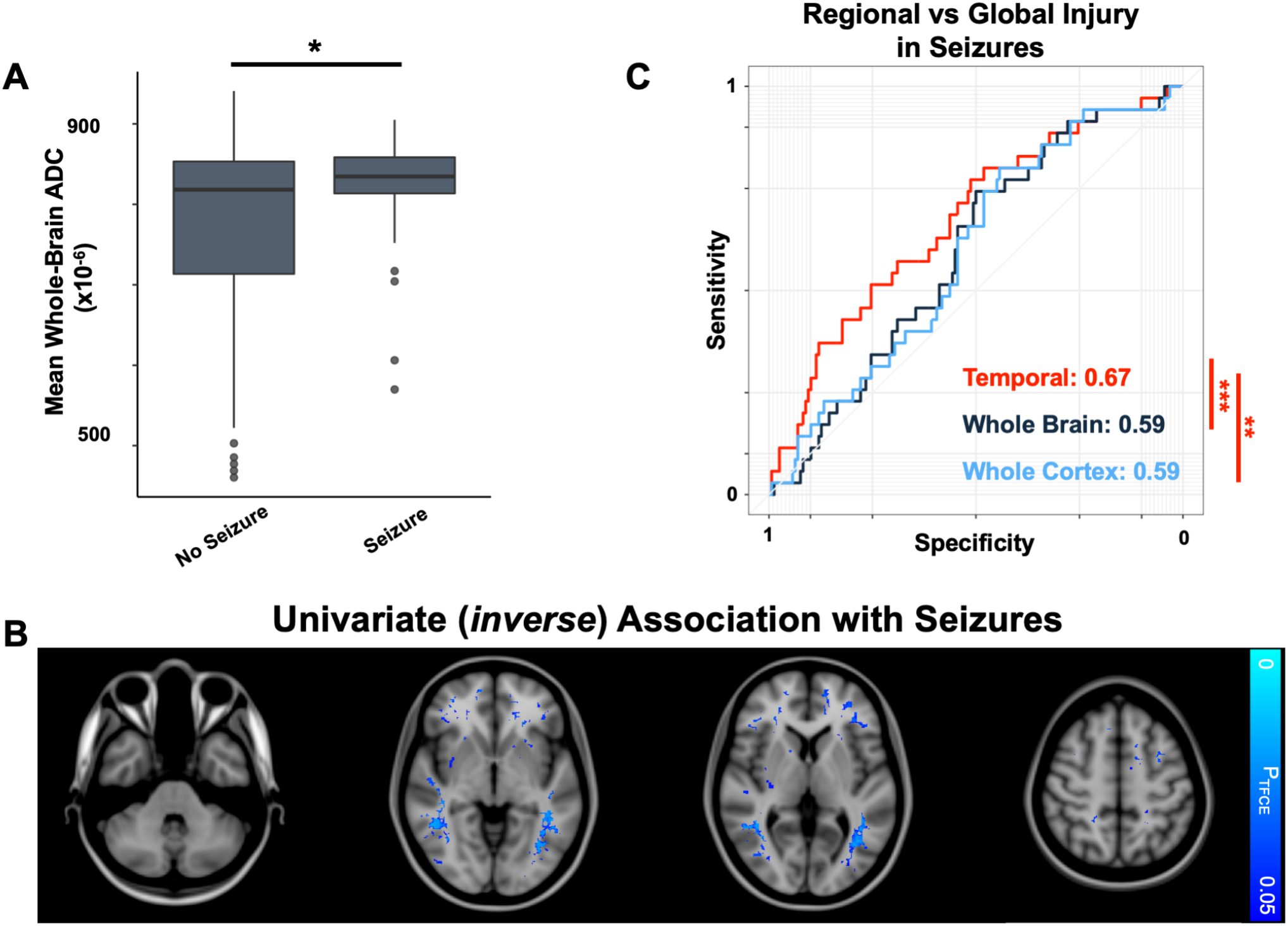
Regional ADC is lower in patients *without* seizures after cardiac arrest. (**A**) Patients without seizures have a lower mean whole-brain ADC compared to those with seizures. **(B)** A TFCE-corrected map of brain voxels with lower ADC in patients without compared to with seizures, illustrating peak differences in the lateral temporal white matter. **(C)** Temporal lobe ADC (red) is superior at classifying patients with versus without seizures compared with whole-brain (dark blue) and whole-cortex (light blue) ADC. AUROC values provided adjacent to legend. *** = *P* < 0.0005, ** = *P* < 0.005, * = *P* < 0.05.

### Seizures: global versus regional

No individual ROIs were associated with seizures using lasso regression. Given that the voxel-wise seizure-association maps highlighted the temporal regions, we explicitly tested whether mean temporal lobe ADC values or whole-brain measures of ADC could better distinguish patients with from those without seizures. Temporal lobe ADC had a greater AUROC (0.67 [0.57, 0.77]) for identifying patients with seizures than either whole-brain (0.59 [0.50, 0.69], *Z* = 3.7, *P* < 0.001) or whole-cortex ADC (0.59 [0.49, 0.69], *Z* = 3.1, *P* = 0.002, Figure 4C). Temporal lobe ADC had a greater AUROC than whole-brain and whole-cortical ADC in 99.1% of 1000 samples of 50% of the data, confirming that these findings were not driven by outlier patients.

## Discussion

Here, we demonstrate that cardiac arrest produces a posterior-predominant anoxic injury pattern. While DoC after cardiac arrest is associated with widespread ABI, it is most strongly associated with the severity of occipital injury. We also find that background suppression is a non-specific marker of overall ABI severity. Seizures are associated with less severe ABI, and lateral temporal anoxia is specifically associated with a lower incidence of seizures.

### Topography of diffusion abnormalities after cardiac arrest

Diminished ADC after cardiac arrest occurs across a range of brain regions. Why adjacent regions are affected to different extents (medial vs lateral thalamus, putamen vs caudate, occipital vs posterior frontal) remains unknown, but may relate to regional differences in metabolic demand^44^ and perfusion^1, 45^.

Primary visual cortex has been theorized to have the highest resting metabolic rate in the brain^44^, which could explain the occipital sensitivity to anoxia, but recent work has disputed this^46^. The posterior predominance seen here is also seen in other neurologic conditions. Posterior reversible encephalopathy syndrome (PRES) is thought to be caused by a diminished autoregulatory reserve of the posterior cerebral circulation^47^. We postulate that PRES and ABI may represent failures at opposite ends of the cerebral autoregulatory curve: a failure of flow limitation at high blood pressures in PRES and a failure to maintain perfusion with low blood pressures in cardiac arrest.

Though prior literature contends that the hippocampus may be particularly vulnerable to anoxia^3^, we did not identify preferential ABI in the hippocampus. However, it possible that these clinical diffusion weighted scans, which poorly discriminate mesial temporal structures^48^, had insufficient resolution to fully assess hippocampal ABI.

### DoC

We found that DoC after cardiac arrest is better predicted by occipital ABI than by the global burden of injury – consistent with theories that posit a posterior predominance of structures important for consciousness^49^. As such, injury to posterior cortical structures may carry more clinical and prognostic value than whole-brain measures of ABI.

The increased diffusivity (ADC) that we observed in patients who recovered consciousness after cardiac arrest has been previously reported^19, 50^, and may reflect mild vasogenic edema^51, 52^. This finding may also reflect unmeasured, fundamental differences between the cardiac arrest and control populations (e.g., intubation, critical illness). Whether this increased diffusivity results in permanent white matter alterations and underlies some of the persistent cognitive complaints recorded in survivors should be investigated^53^

### Background suppression

While occipital and cerebellar ABI showed the strongest regional associations, they were not superior to whole-brain summary measures at predicting background suppression. How such injury results in background suppression remains unknown. Though the thalamus is classically thought to produce the oscillatory signals underlying the continuous EEG background^54^, autopsy studies have found cortical and cerebellar injury more frequently than thalamic injury in patients with background suppression after cardiac arrest, consistent with our findings^3, 55^. We found background suppression to be a relatively non-specific marker of ABI severity. Indeed, other groups have postulated that background suppression may even be neuroprotective after cardiac arrest^11^, representing a functional adaptation of neurons to a metabolic crisis.

### Post-anoxic seizures

Reduced whole-brain ADC values, and reduced temporal lobe ADC values in particular, were associated with a decreased prevalence of seizures. Widespread anoxic injury may leave behind too few intact neuronal elements to generate the synchronized neuronal activity required for seizures. Injury to the lateral temporal lobes specifically may mimic a functional temporal lobectomy, which is known to inhibit seizures^56^. Alternatively, uncontrolled seizures may cause vasogenic temporal lobe edema, which might elevate ADC signal. The possible association between temporal lobe lesions and seizure inhibition may have broader therapeutic implications, warranting further study.

## Limitations

Diffusion-weighted MRI is limited in evaluating brain regions near the skull base^57^, and thus our sensitivity for ABI in these regions may have been similarly limited. Furthermore, while including diffusion acquisitions across multiple scanner types and field strengths permitted a larger sample size, such heterogeneity may have added variance to our data and reduced our power to identify regional associations.

EEG continuity and seizures were not systematically classified prior to 2013, and variability in documentation may have degraded the quality of EEG variables. Furthermore, we did not have adequate power to classify seizure subtypes. It is possible that different types of cerebral lesions predispose to different types of seizures (e.g., myoclonic seizures).

Given that we did not assess why and how life sustaining therapy was withdrawn, it is possible that some patients with DoC or persistent background suppression may have had an intact neural substrate for recovery, but died before such a recovery could be realized. Misclassifications such as these would have reduced our power to identify specific associations with regional injury. Future research with stricter methods for controlling for mortality may further refine these associations.

Finally, while we demonstrate that these findings are robust to outliers within our dataset, it is important to acknowledge the limited generalizability and possible confounds that characterize any retrospective, single-center study.

## Conclusions

In a large retrospective dataset of unresponsive patients after cardiac arrest, we identified the brain-wide distribution of ABI as measured with diffusion MRI, finding more involvement in posterior cortical regions. DoC was associated with anoxic injury to a broad range of cortical and subcortical regions but could be classified most accurately by the severity of occipital injury. Background suppression, a common EEG finding after cardiac arrest, was also a non-specific marker of overall ABI severity and was not preferentially associated with anoxic injury to a particular region. Finally, seizures were less frequent in patients with severe ABI, and especially after anoxia to the lateral temporal lobes. In total, our results suggest that the regional pattern of anoxic brain injury after cardiac arrest may hold importance for prognosis and salient clinical sequelae.

## Data Availability

available upon reasonable request to corresponding author.

## Supplementary material

Supplementary figures referenced in the text are provided in the accompanying pdf.

## Abbreviations

ABI: Anoxic Brain Injury
ADC: Apparent Diffusion Coefficient
DoC: Disorder of Consciousness
ROI: Region of Interest
TTM: Targeted Temperature Management
VLSM: Voxel Lesion Symptom Mapping

## SUPPLEMENTARY MATERIAL

**Supplementary Figure 1:**
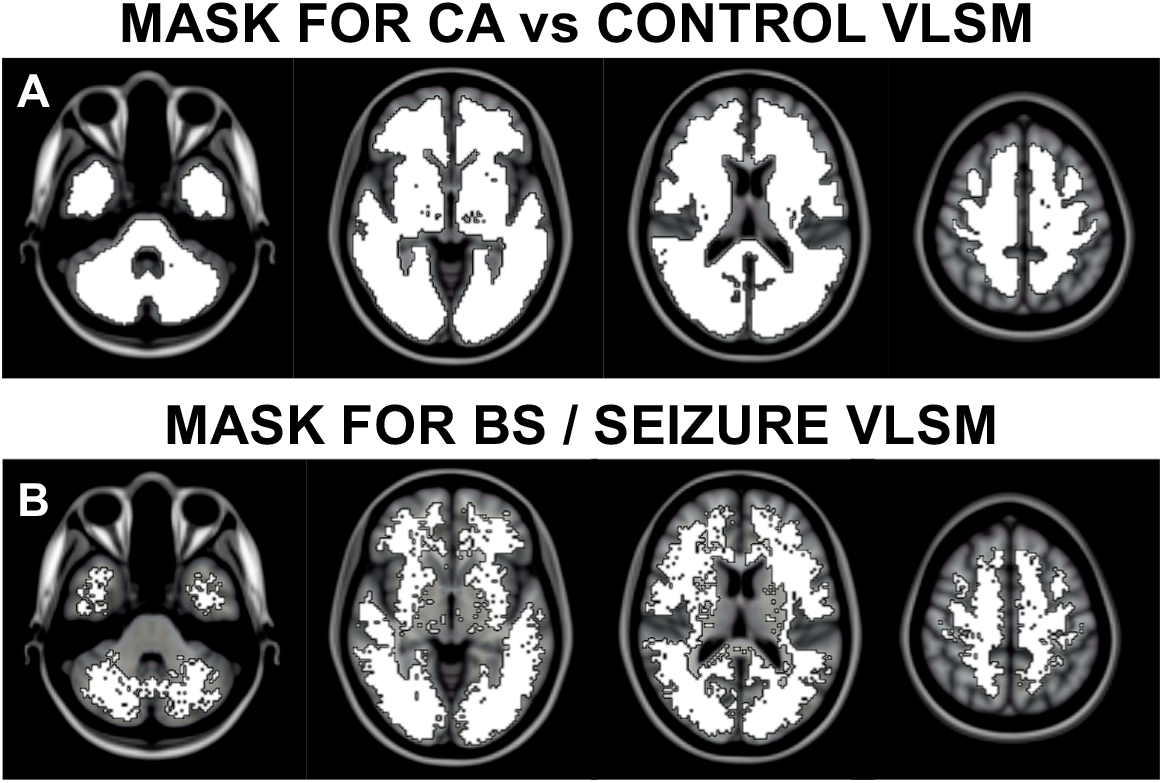
VLSM Masks. (A) Voxels included for the cardiac arrest (CA) versus control VLSM analysis. This mask excluded voxels with an ADC value of 0 in more than 20% of patients. (B) Voxels included in this the VLSM analysis for background suppression (BS) and seizures. Includes all significant voxels (P_TFCE_ < 0.05) from the DoC vs Recovered VLSM.

**Supplementary Figure 2:**
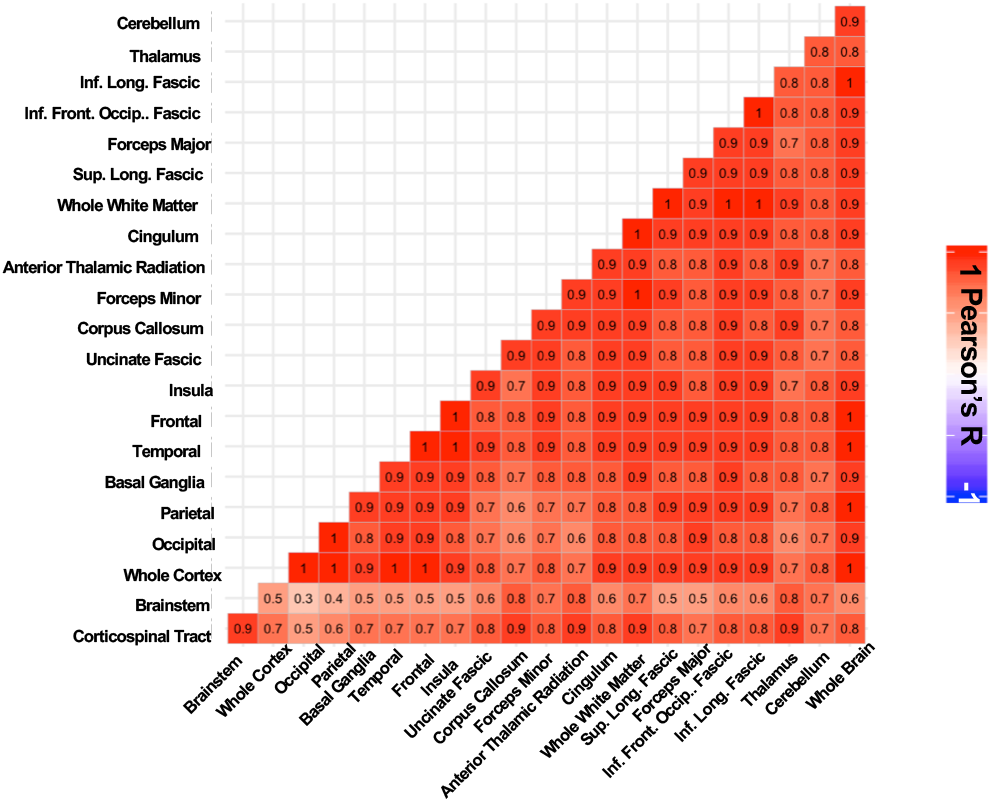
Mean regional ADC measurements are highly correlated. Correlation matrix (Pearson’s R) of the mean ADC signal in each ROI.

**Supplementary Figure 3:**
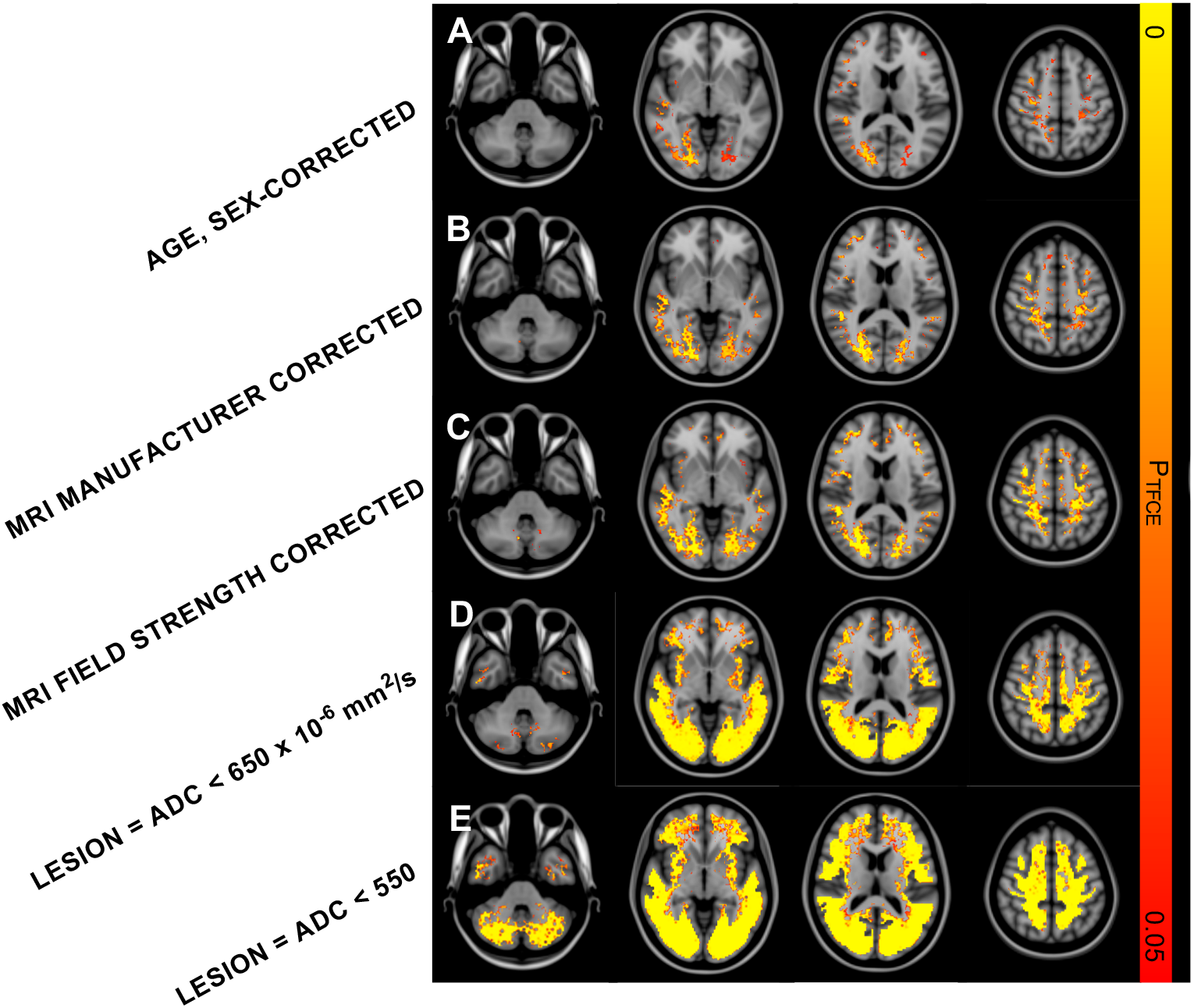
Cardiac Arrest vs Control: sensitivity analyses. VLSM results comparing ADC maps between cardiac arrests and controls. Significant voxels (P_TFCE_ < 0.05) for each analysis are shown. (A) Adjusted for mean-centered age and sex. (B) Adjusted for MRI manufacturer (Siemens vs GE). (C)Adjusted for field-strength (1.5T vs 3T). (D) VLSM after converting each subject’s ADC map into a lesion map by binarizing all ADC values less than 650 × 10^−6^ mm^2^/s or (E) 550 × 10^−6^ mm^2^/s.

**Supplementary Figure 4:**
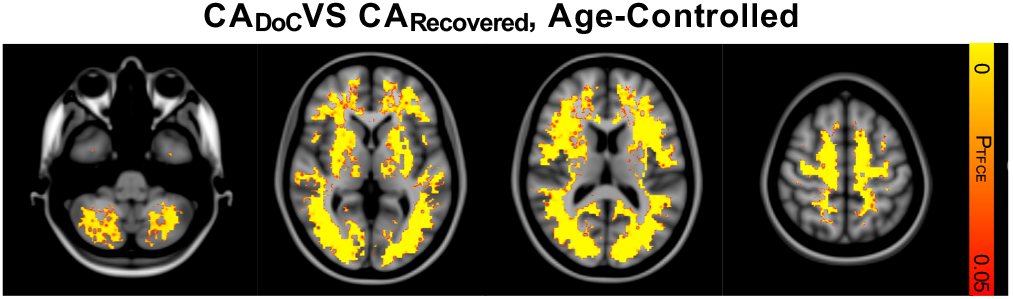
Differences between CA_DoC_ and CA_Recovered_ persist after controlling for age. (A) Significant voxels (P_TFCE_ < 0.05) associated with DoC after cardiac arrest, controlling for mean-centered age.

**Supplementary Figure 5:**
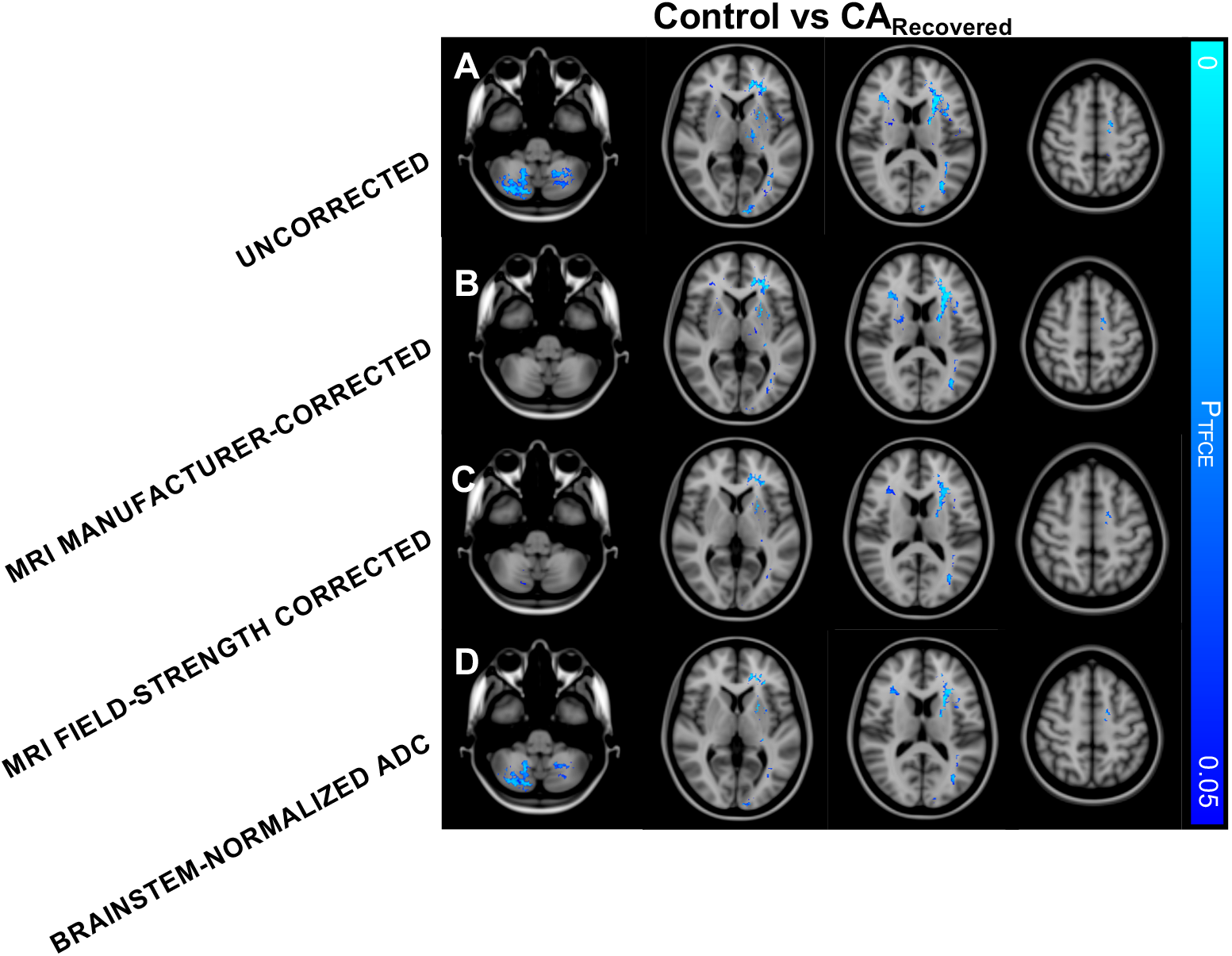
ADC distributions in controls compared to CA_Recovered_. (A) Voxels with significantly different ADC distributions between CARecovered patients and controls (P_TFCE_ < 0.05). All significant voxels showed lower ADC in controls compared to CARecovered. A similar voxelwise distribution was observed after controlling for the nuisance covariates of (B) MRI manufacturer (GE vs Siemens), (C) MRI field strength (1.5T vs 3T) and (D) normalizing each subject’s ADC map to the mean ADC within the brainstem.

**Supplementary Figure 6:**
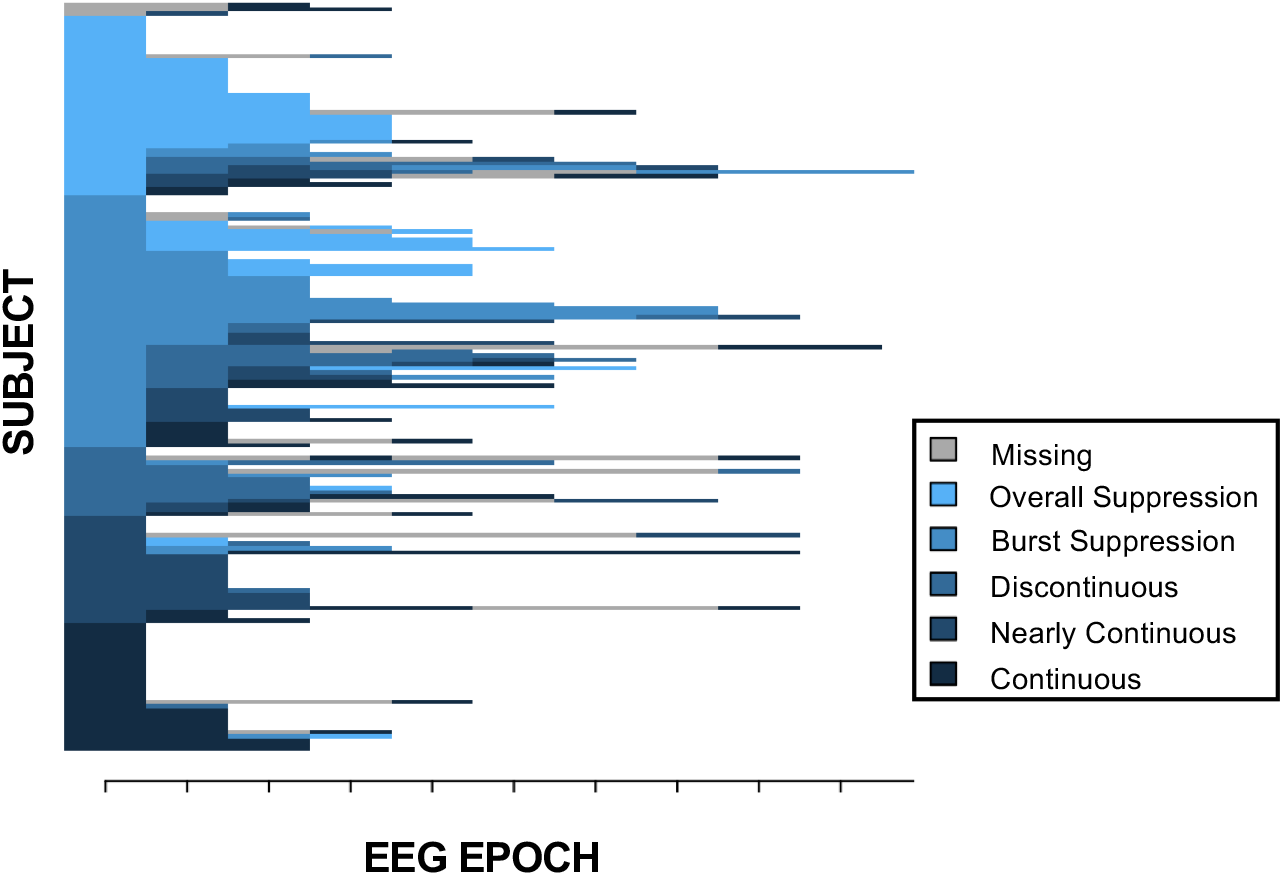
Individual subject EEG background continuity trajectories. Individual subject EEG background continuities are shown for each post-arrest 24 hour EEG epoch.

**Supplementary Figure 7:**
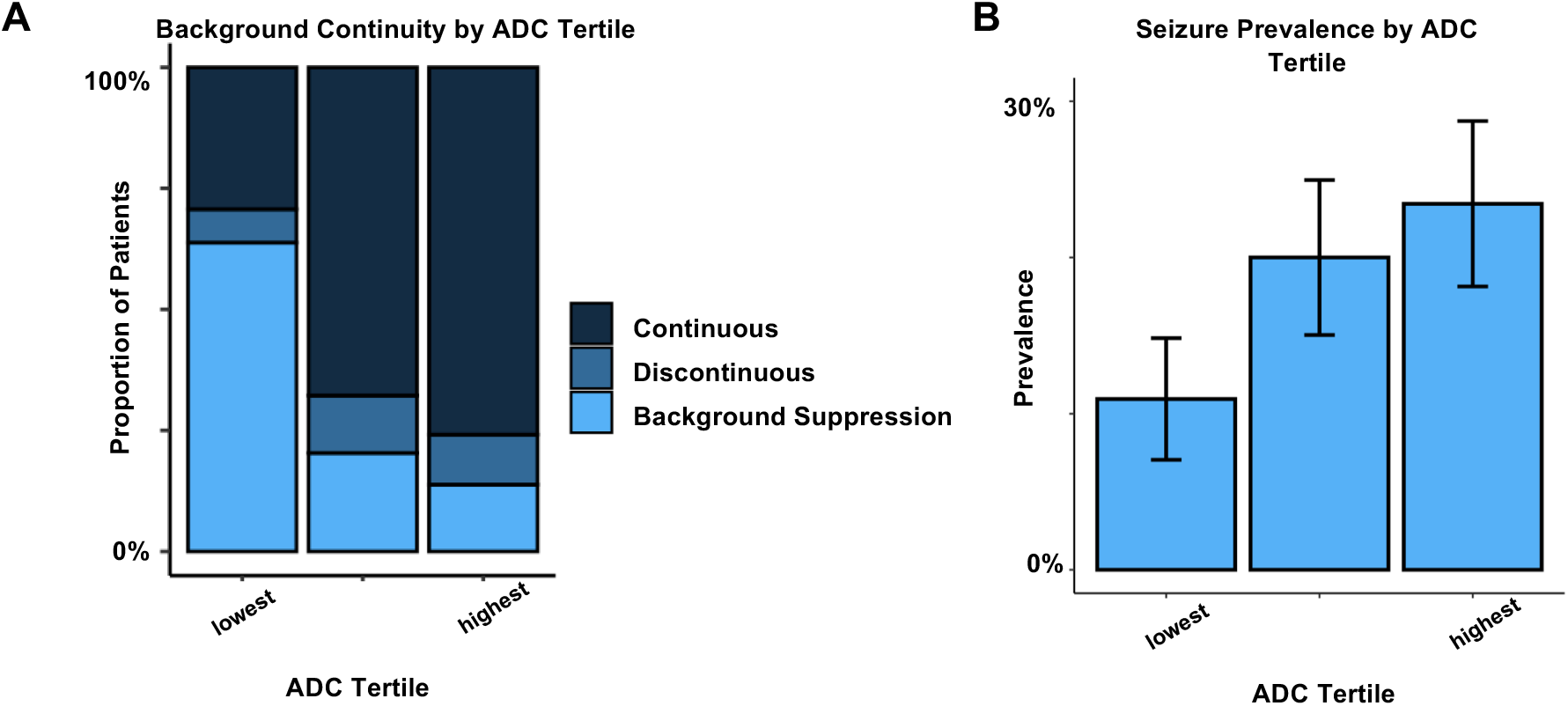
Prevalence of EEG features by ADC tertile. (A) Proportion of patients with each class of best-achieved EEG background at each tertile of whole-brain mean ADC values [lowest: < 792 ×10^−6^ mm^2^/s, middle: 792-844, highest > 844]. (B) Proportion of patients with seizures at each tertile of whole-brain mean ADC values. Error bars represent standard error of the estimated proportion.

**Supplementary Figure 8:**
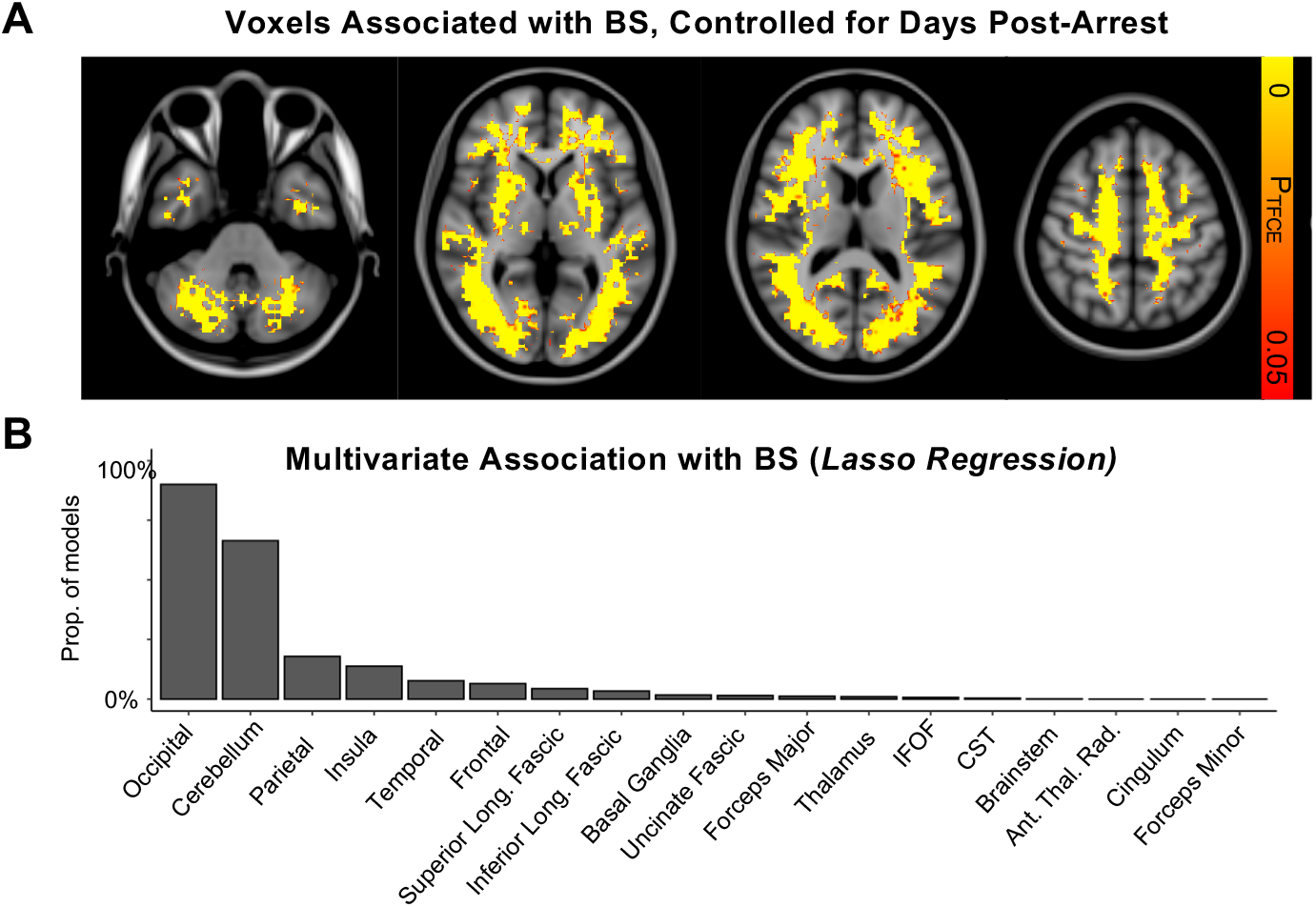
Robustness of regional associations with background suppression (BS) (A) Voxels significantly associated with the presence of persistent background suppression after cardiac arrest, controlling for the nuisance covariate of mean-centered days between arrest and MRI acquisition (P_TFCE_ < 0.05). (B) Proportion of 1000 split-half iterations in which each anatomical ROI was included in the lasso regression with a non-zero coefficient. Abbreviations: IFOF = Inferior Fronto-Occipital Fasciculus, CST= Corticospinal Tract.

